# Cross-phenotype relationship between opioid use disorder and suicide attempts: new evidence from polygenic association and Mendelian randomization analyses

**DOI:** 10.1101/2022.11.03.22281876

**Authors:** Yunqi Huang, Dongru Chen, Albert M. Levin, Brian K. Ahmedani, Cathrine Frank, Miaoxin Li, Qiang Wang, Hongsheng Gui, Pak C. Sham

## Abstract

**IMPORTANCE:** Clinical epidemiological studies have found high rates of comorbidity between suicide attempts (SA) and opioid use disorder (OUD). However, the patterns of correlation and causation between them are still not clear due to psychiatric confounding.

**OBJECTIVE:** To investigate the pairwise associations and interrogate the potential bidirectional relationship between OUD and SA using genetically based methods.

**DESIGN, SETTING, AND PARTICIPANTS:** We utilized raw phenotypes and genotypes from UK Biobank, and summary statistics from Million Veteran Program, Psychiatric Genomic Consortium, iPSYCH, and International Suicide Genetics Consortium. Statistical genetics tools were used to perform epidemiological association, genetic correlation, polygenic risk score prediction, and Mendelian randomizations (MR). Analyses were conducted to examine the OUD-SA relationship with and without controlling for psychiatric disease status (e.g., major depressive disorder [MDD]).

**MAIN OUTCOMES AND MEASURES:** OUD and SA with or without major psychiatric disorders (schizophrenia, bipolar disorder, major depressive disorder, and alcohol use disorder).

**RESULTS:** Strong correlations between OUD and SA were observed at both phenotypic level (overall samples [OR=2.94, *P* =1.59 ×10^−14^]; non-psychiatric subgroup [OR=2.15, *P* =1.07 ×10^−3^]) and genetic level (r^2^=0.4 and 0.5 with or without conditioning on MDD). The higher genetic susceptibility to SA can increase the polygenic risk of OUD (OR=1.08, false discovery rate [FDR] =1.71 ×10^−3^), while the higher susceptibility to OUD can also increase the risk of SA (OR=1.09, FDR =1.73 ×10^−6^). However, predictive abilities for both were much weakened after controlling for influence of psychiatric diseases. A combination of different MR analyses suggested a possible causal association from SA to OUD (2-sample univariable MR: OR=1.14, *P* = 0.001; multivariable MR: OR=1.08, *P* = 0.001).

**CONCLUSIONS AND RELEVANCE:** This study provided new genetic evidence underlying the strong OUD-SA comorbidity. While controlling for the influence of psychiatric diseases, there is still some clue on possible causal association between SA genetic liability and the risk for OUD. Future prevention strategy for each phenotype needs to take into consideration of screening for the other one.

**Key Points:** *Question:* Does opioid use disorder (OUD) have a potentially causal role in the risk for suicide attempts (SA), or vice versa?

*Findings:* In this observational study with raw data from >150,000 UK Biobank samples and genome-wide association summary statistics from >600,000 individuals, positive phenotypic and genetic associations were observed between OUD and SA, whether or not controlling for major psychiatric disorders. Comprehensive Mendelian randomization analyses (one-sample and two-sample) suggested the genetic liability for SA was associated with increased risk for OUD. However, they were still underpowered to reveal the putative causal association from OUD to SA.

*Meaning:* This genomics-based study supports a strong genetic association underlying the OUD-SA comorbidity. Though both phenotypes are intertwined with other psychiatric disorders, there also exists an independent bidirectional relationship between OUD and SA.

## Introduction

Opioids are among the world’s oldest known psychoactive drugs for thousands of years, and widely used for medicinal and recreational purposes.^1^ Fentanyl, tramadol, hydromorphone and morphine, the most commonly prescribed opioids, are used in acute and chronic pain management, and non-pain conditions such as multiple sclerosis.^2, 3^ Although the total number and rate of opioid prescription dispensed have declined in the last decade, the incidence of opioid use disorder (OUD) and rate of opioid overdose death are both increasing.^4, 5^ Over 40 million people suffer from opioid misuse or OUD globally.^1, 6, 7^ In the United States, it is estimated that >10 million people misused opioid, >2 million had an OUD, and ∼50,000 died from opioid overdose annually.^8^ However, not everyone who has opioid exposure (OE) extra-medically develops OUD,^9–11^ and the underlying biological mechanisms of developing OUD are to be uncovered.

Suicidality is another major public health concern that accounts for death and disability worldwide with an increased rate in recent decades^12, 13^. Suicide attempts (SA), defined as non-fatal self-injurious behaviour with the intent to die, has been estimated to occur about 10-20 times more frequently than actual suicide and is a major source of disability, reduced quality of life, and social and economic burden.^14^ Known risk factors for SA are mental health (MH) conditions, such as major depressive disorder (MDD), bipolar disorder (BD), schizophrenia (SCZ) and their related clinical variables.^15–17^ Opioid drugs are one of the mostly used means for committing suicide.

Clinical epidemiological studies have found high rates of comorbidity between these two behavioral traits,^18–20^ especially withdrawal from or forced tapering states.^21^ As a reflection of their interconnected relationship, risk of suicidal ideation and behaviour has been found markedly increased in people with OUD, with highest risk for more severe outcomes.^22–24^ Also, these two public health crises are intertwined at multiple levels. People with a range of mental health conditions, social and environmental factors are at high risk for both disorders.^25–27^ Chronic pain, one of the reasons of opioid medical use, is associated with increased risk of mood or anxiety disorder^28^ and SA.^29^ However, due to confounding factors, it is challenging to explain OUD-SA comorbidity with epidemiology data.

In fact, both SA^30^ and OUD^31^ are genetically heritable (heritability at 0.55 and 0.34, respectively), and have been investigated through genome-wide association studies (GWAS). The largest-scale GWAS on OUD, performed by Million Veteran Project (MVP) researchers, reidentified the *OPRM1* gene as the most replicable genetic locus.^31^ A recent GWAS on SA that was performed by International Suicide Genetics Consortium (ISGC), has assembled almost all available SA datasets in the field.^30^ Besides identifying novel disease genes for each phenotype, these updated genomic data also provide us new resource for interrogating cross-phenotype relationships, for instance, through polygenic risk score (PRS) prediction and Mendelian randomization (MR)^32^ analysis. These two approaches are complementary to each other and can rigorously assess the evidence regarding association and causality when combined.^33^ This is also an opportunity to quantify their genetic correlation in the context of other psychiatric diseases, and then help to dissect their phenotypic correlation into nature and nurture parts.

Up to date, there is still no valid longitudinal cohort or clinical trial to reveal the temporal pattern and mechanism behind OUD and SA co-occurrence. Here we combined UK Biobank (UKB) phenome-genome data and published GWAS summary statistics to investigate their multi-layer relationships. The statistical genetics approaches were tailored for the data and adopted to reveal the possibility and degree of bidirectional association between OUD and SA. Our study also controlled for potential confounders of their psychopathology by conditional and stratified models. This new evidence may help us design more efficient strategy for promoting behavioural health in future.^26, 27, 34^

## Materials and Methods

### Study design

Genomic data and statistical genetics analyses provide a new solution to disentangle the comorbidity between OUD and SA. As shown in **eTable 1**, we have collected both primary data (UK Biobank) and secondary data (GWAS summary statistics) and designed an integrated analytical pipeline to investigate their relationship (**eFigure 1**).

**Table 1.** Descriptive statistics for SA and OUD assessed in UKB. Total size 3591 116674 2169 50543

| Characteristic | SA |  |  | OUD |  |  |
| --- | --- | --- | --- | --- | --- | --- |
|  | Cases | Controls | P | Cases | Controls | P |
| Total size | 3591 | 116674 |  | 2169 | 50543 |  |
| # Male (%) | 1290 (35.9) | 52299 (44.8) | $<2.2 \times 10^{-16}$ | 817 (37.7) | 21024 (41.6) | $3.0 \times 10^{-4}$ |
| Mean age (SD) | 53.9 | 56.4 | $<2.2 \times 10^{-16}$ | 59 | 58.6 | 0.026 |
| # MH diseases (%)<sup>*</sup> | 1242 (34.6) | 8775 (7.5) | $<2.2 \times 10^{-16}$ | 826 (38.1) | 9922 (19.6) | $<2.2 \times 10^{-16}$ |
| # OUD (%) | 67 (1.9) | 338 (0.3) | $<2.2 \times 10^{-16}$ | 2169 (100.0) | 0 (0.0) | NA<sup>^</sup> |
| # SA (%) | 3591 (100.0) | 0 (0.0) | NA<sup>^</sup> | 67 (3.1) | 775 (1.5) | $2.8 \times 10^{-8}$ |

<sup>\*</sup> Mental health (MH) diseases were defined by ICD-9/10 codes for depression, anxiety, stress-related disorder, substance misuse (excluding OUD), and psychotic disorders.
<sup>^</sup> NA means not available as the test is not valid when certain cell count is 0.
SD shorts for standard deviation.

**Figure 1.**
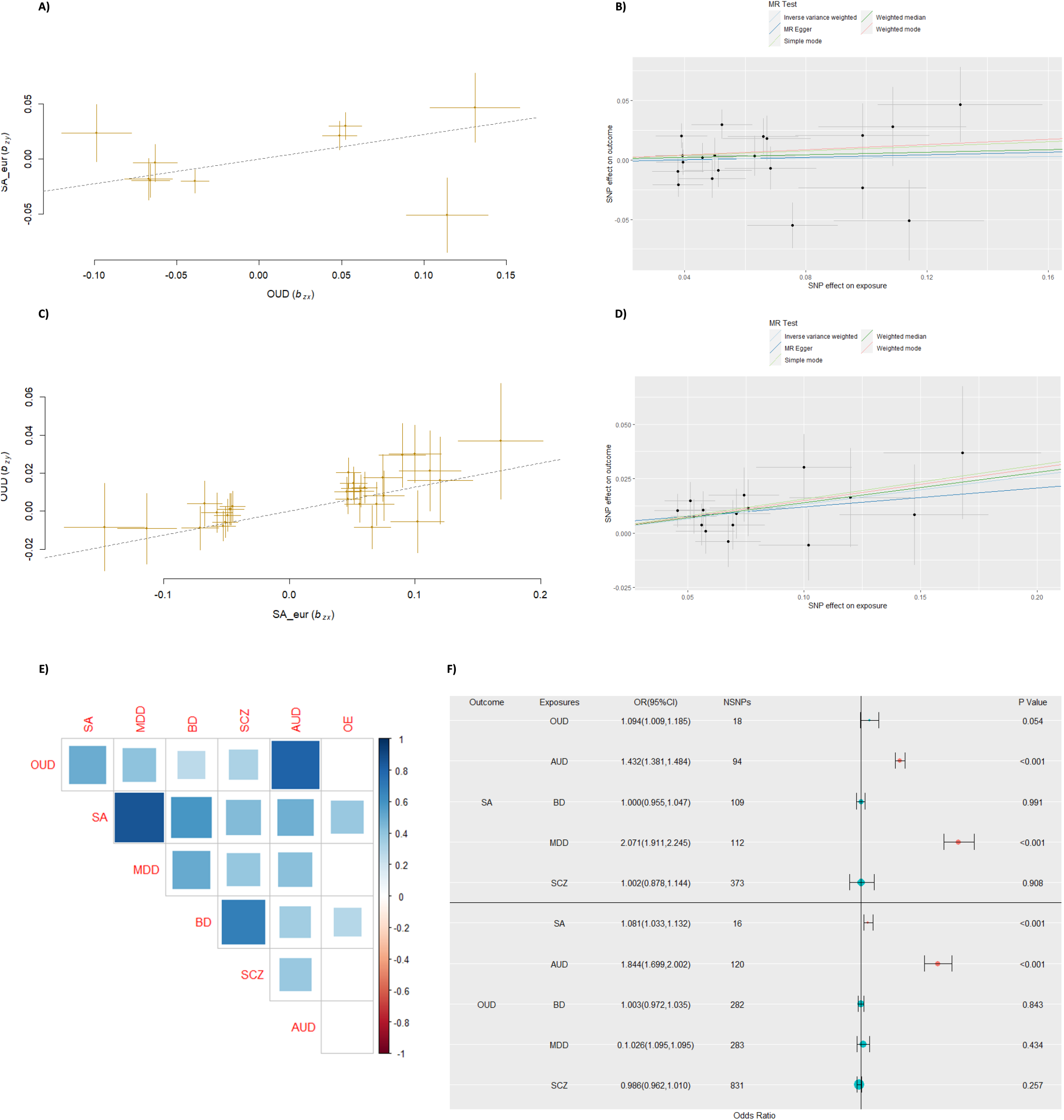
Bi-directional two-sample Mendelian randomization results between OUD and SA. A) Results of GSMR analysis from OUD to SA. Yellow crosses: 9 SNPs included in GSMR. B) Results of IVW, Weighted median, MR-Egger, Weighted mode and simple mode from OUD to SA. Black crosses: 20 SNPs included in the analyses. C) Results of GSMR analysis from SA to OUD. Yellow crosses: 29 SNPs included in GSMR. D) Results of IVW, Weighted median, MR-Egger, Weighted mode and simple mode from SA to OUD. Black crosses: 16 SNPs included in the analyses. E) Genetic correlations across different phenotypes included in multivariable Mendelian Randomization. F) Odds ratios (ORs) and 95% confidence intervals (CIs) for the effect of SA liability on OUD and vice versa, estimated using the multivariable Mendelian randomization (MR) IVW approach. The vertical line means OR = 1. Red points represent significant causal effects, and green points represent unsignificant associations. The larger point size is, the more significant SNPs of the traits were included in the analyses.

### UK Biobank

Definition of OUD and SA phenotypes (binary status), and other comorbid psychiatric traits were through mental health questionnaire (MHQ) and electronic health records in UKB (**eTable 2**). Processing of genotype data followed the recommendation in the original report.^35^ Detailed criteria and quality control steps are included in the Supplementary. Particularly, we used self-reported ancestry, principal component (PC) analysis and kinship relatedness to select European-descent unrelated individuals.^36, 37^ Number of PC to be adjusted for downstream association was determined by their eigenvalues (**eFigure 2)**. The UKB received ethical approval from the North West–Haydock Research Ethics Committee (REC reference 11/NW/0382), and had appropriate informed consent from all its participants. The current study was conducted under application No. 15422 and 86920.

**Table 2.**
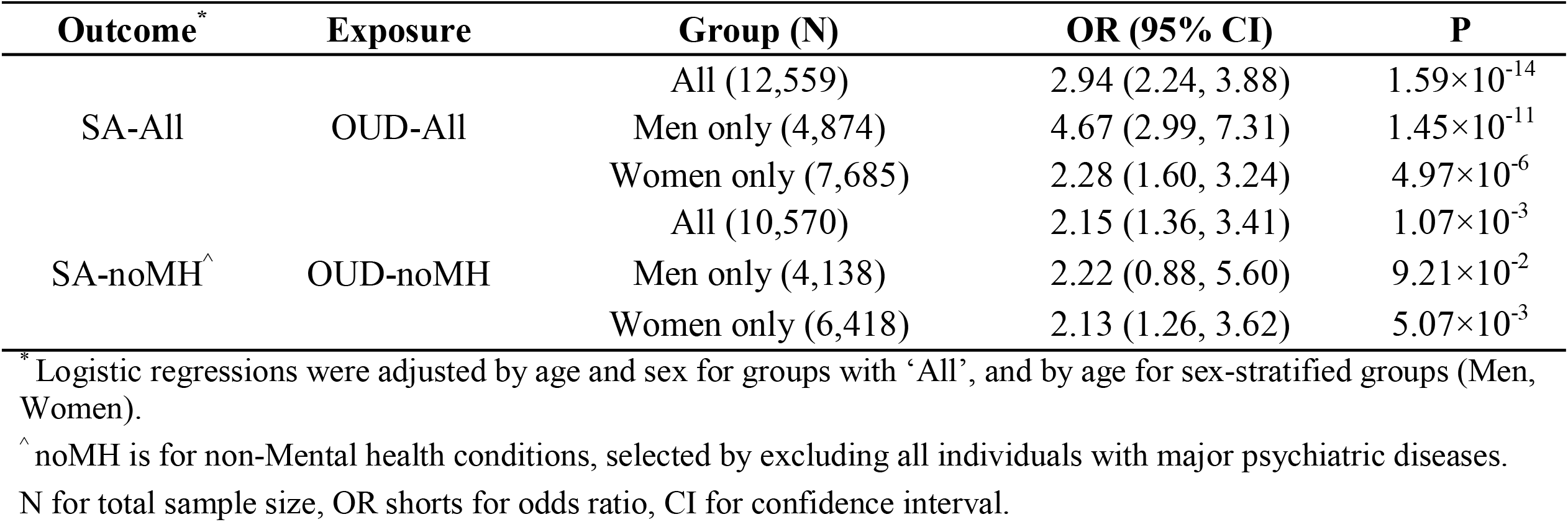
Observational association between SA and OUD phenotypes in UKB.

### GWAS summary statistics

GWAS summary statistics for SA were collected from two resources: 1) overall ISGC data repository (26,590 cases, 492,022 controls; with UKB),^30^ the largest GWAS focusing on SA; and 2) a Danish Integrative Psychiatric Research (iPSYCH) study (6,024 cases, 44,240 controls; without UKB)^38^ as independent verification. Genetic variants presented in both datasets showing no heterogeneity were retained. GWAS summary statistics for OUD (10,544 cases, 72,163 controls; without UKB) and alcohol use disorder (AUD) was accessed through NCBI dbGaP for MVP (phs001672.v6.p1).^39^ GWAS summary statistics for OE, MDD, BD and SCZ were downloaded directly from Psychiatric Genomics Consortium (PGC) website.^40, 41^ Additional harmonization steps were included in the Supplement.

### Observational association in UKB

Descriptive statistics between groups were compared and test for difference by independent t-test or chi-square test. Conditional logistic regression was used to examine the observational association between OUD status and SA outcomes in all eligible subjects (OUD-All and SA-All) and those without mental health problems (OUD-nonMH and SA-nonMH) among UKB participants. Model was adjusted by age, gender, and whether diagnosed with mental health disorders or not. The age and sex of participants were baseline characteristics determined at recruitment. These conditional models were run in all individuals, and in males and females separately.

### Polygenic risk score association

Published GWAS for MVP OUD^39^ or iPSYCH SA^38^ was used as training dataset, and UKB data as target dataset. ISGC SA data was not used in PRS analysis since it already included UKB. Training dataset included GWAS summary statistics with and without adjustment of major mental health diseases (iPSYCH M2 and M1; MVP OUD|MDD and OUD). HapMap3 CEU markers^42^ was referred in this analysis. PRSice-2^43^ was used to construct PRS scores in UKB targeted dataset at different *P* thresholds (5×10^−8^, 10^−7^, 10^−6^, 10^−5^, 10^4^, .001, .01, .05, 0.1, 0.2, 0.3, 0.4, 0.5, and 1). To avoid systematic error from population stratification,^44^ a principal component analysis (PCA) on the resulting PRS scores was performed and the resulting PCs were used in subsequent association analysis.^45^ Standardized PRS was generated using the entire distribution and associated with binary phenotype (OUD or SA) by logistic regression, adjusted for age, sex, PC1-PC4, and array type.

### Univariable Mendelian Randomization

Only OUD and SA were included in this type of MR. First, bidirectional one-sample MR (1SMR) analyses were performed on UKB using individual-level genotyping data matched with corresponding phenotypes in ‘All’ or in those ‘nonMH’ samples. Second, bi-directional two-sample MR (2SMR) was then conducted by inverse variance–weighted (IVW) method (as primary) and 3 other companion methods (weighted median, MR-Egger regression, and GSMR; as secondary), imbedded in TwoSampleMR and MendelianRandomisation R packages (R version 4.0.2) and GSMR software^46^. Pleiotropy or heterogeneity test was performed with MR-Egger regression (intercept), MR-PRESSO, or HEIDI test in GSMR, respectively. Several sensitivity analyses were included to further evaluate the influence of different instrumental SNPs, overlapping sample, and confounding factors on 2SMR estimated effects (detail in Supplement). MR power calculation was performed using mRnd.^47^

### Pairwise genetic correlation

To examine how other psychiatric exposures may serve as potential confounders, we firstly applied linkage disequilibrium score regression (LDSC)^48^ to estimate the genetic correlation between OUD, SA and related MH phenotypes. The Bonferroni method was used to correct the *P* values returned from LDSC. Only those having significant genetic correlation with OUD or SA were included in final multivariable MR analyses.

### Multivariable Mendelian Randomization

Multivariable MR (MVMR) analyses were lastly performed using GWAS summary dataset from OUD, SA, and their correlated psychiatric traits. The same criterion as univariable MR was adopted to select instrumental SNPs for each exposure, covariate, or outcome. We used the MVMR extension of the IVW method to estimate the effect of each exposure trait and whether there existed mediating effects from its covariates. Multivariable median method and MR-Egger method were also included as sensitivity tests. Forest plots were used to present the MVMR fitting.

## Results

### Observational association between OUD and SA

When examined individually, male gender, younger age, psychiatric diseases affection, and OUD are all risk factors for SA; as a comparison, male gender, older age, psychiatric diseases affection, and SA are risk factors for OUD (**Table 1** and **eTable 3**). In the conditional logistic models (**Table 2**), patients diagnosed with OUD were associated with higher likelihood of SA in overall sample (OR =2.94; 95% CI, 2.24-3.88; *P* =1.59×10^−14^). After limiting to those without major mental health disorders (nonMH), their phenotypic correlation is still significant (OR =2.15; 95% CI, 1.36-3.41; *P* =1.07×10^−3^). For the gender-specific analysis, OUD status presented positive association with SA in both gender groups but had stronger effect in males than in females (OR =4.67 vs 2.28 in overall, and 2.22 vs 2.13 in non-MH).

**Table 3.**
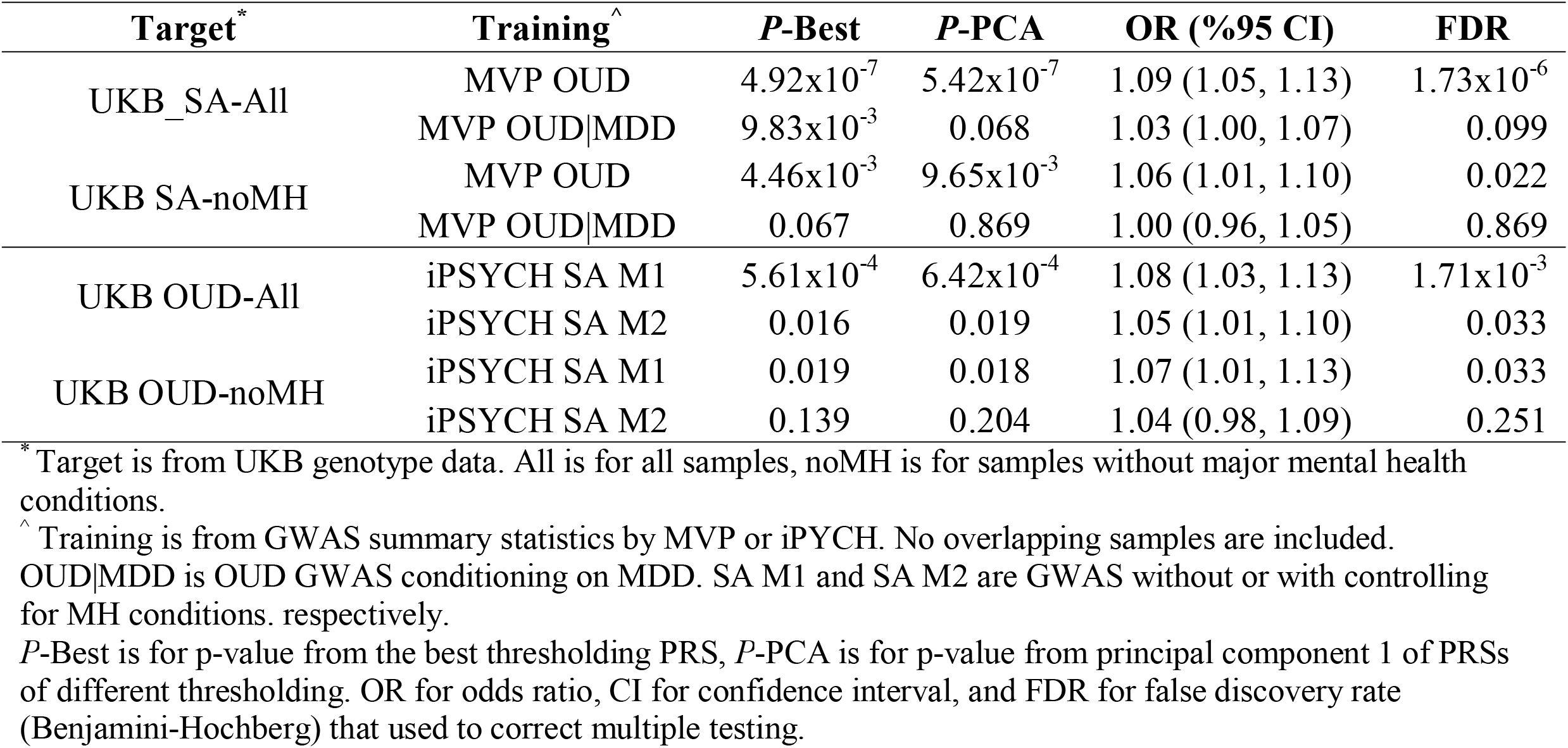
Polygenic risk score association between OUD and SA.

### Polygenic risk score association between OUD and SA

As shown in **Table 3**, the PRS for MVP OUD significantly predicted SA status in UKB, among all and non-MH samples (OR =1.09 or 1.06, respectively; FDR < .05). Meanwhile, the PRS for iPSYCH SA was also positively associated with OUD status in UKB for both settings (OR=1.08 or 1.07, respectively; FDR <.05). When using GWAS with adjustment of major psychiatric diseases as training data, the PRS association was still significant for the pair of iPsych SA (M2) and UKB OUD-All (OR =1.05, FDR <.05); but the effects for other three OUD-SA pairs were much weakened and not significant anymore.

### Univariable MR-based association between OUD and SA

Using PRS as an instrumental variable, 1SMR in UKB data did not identify any significant causal association between OUD and SA (**eTable 4**). In the extended 2SMR result (**Table 4)**, it showed a significant causal effect of the genetic risk of SA on OUD (IVW: OR =1.14, 95%CI: 1.05-1.24, *P* =.001; GSMR: OR =1.14, 95%CI: 1.07-1.21, *P* <.001), surviving Bonferroni correction (0.05/8). Three validity tests did not find any violation of MR assumptions for SA-OUD analysis (all *P* >0.05). However, no consistent evidence of a reverse causal effect was found when investigating the direction from OUD to SA (IVW: OR =1.02, 95%CI: 0.87-1.19, *P* =.828; GSMR: OR =1.25, 95%CI:1.05-1.49, *P* =.014). This may be due to the existence of horizontal pleiotropy (MR-PRESSO global test *P* = .023) and limited MR power (<0.2). The overall distribution of instrumental SNPs’ effects for both 2SMR analyses are shown in **Figure 1A-D**. When re-examining their relationship in additional sensitivity analyses (**eTable 5**), the causal effect of SA liability on OUD is still significant (*P* < .05) after regressing out the genetic risk of MDD from both GWAS datasets. Like 1SMR result, none of these causal associations was identifiable when using much smaller SA GWAS (i.e., iPSYCH cohort) against MVP OUD. More details on model fitting and individual SNP annotation are included in **eTables 6 and eFigures 3-4**.

**Table 4.** Univariable two-sample MR result using MVP-OUD and ISGC-SA datasets.

| Exposure vs Outcome<sup>^</sup> | MR effect estimate |  |  |  |  | MR validity test |  | MR power |  |
| --- | --- | --- | --- | --- | --- | --- | --- | --- | --- |
|  | Method | OR | 95% CI | *P* | nsnp<sup>*</sup> | Method | *P* | F-statistic | Power |
| OD ~ SA | IVW | 1.02 | 0.87-1.19 | 0.828 | 20 | MR-Egger intercept test | 0.868 | 6957.83 | 0.16 |
|  | Weighted median | 1.06 | 0.89-1.26 | 0.538 | 20 | MR-PRESSO global test | 0.023 |  |  |
|  | MR-Egger regression | 1.06 | 0.66-1.70 | 0.207 | 20 | Q statistics heterogeneity test | 0.023 |  |  |
|  | GSMR | 1.25 | 1.05-1.49 | 0.014 | 9 |  |  |  |  |
| SA ~ OD | IVW | 1.14 | 1.05-1.24 | 0.001 | 16 | MR-Egger intercept test | 0.726 | 10695.47 | 1 |
|  | Weighted median | 1.15 | 1.03-1.28 | 0.012 | 16 | MR-PRESSO global test | 0.892 |  |  |
|  | MR-Egger regression | 1.09 | 0.84-1.42 | 0.525 | 16 | Q statistics heterogeneity test | 0.966 |  |  |
|  | GSMR | 1.14 | 1.07-1.21 | <0.001 | 29 |  |  |  |  |

<sup>^</sup> In two-sample MR between MVP and ISGC, overlapping sample is only 0.4%.
<sup>\*</sup> nsnp means number of instrumental SNPs used. More SNP details are provided in TabS6.
OR stands for odds ratio, CI for confidence interval, MR for Mendelian randomization, IVW for inverse-variance weighted, GSMR for generalized summary-data-based Mendelian randomization.

### Genetic correlation with other psychiatric traits

LDSC result (**Figure 1E** and **eTable 7**) showed that OUD and SA had a high genetic correlation (rg =0.50, *P* =1.81×10^−11^). Both traits also presented strong correlations with MDD (OUD: rg =0.41, *P* = 2.40×10^−8^; SA: rg =0.86, *P* =2.51×10^−123^), BD (OUD: rg =0.27, *P* =1.33×10^−6^; SA: rg =0.58, *P* =6.13×10^−81^), SCZ (OUD: rg =0.31, *P* =2.55×10^−10^; SA: rg =0.43, *P* =9.58×10^−56^) and AUD (OUD: rg =0.81, *P* =2.64×10^−39^; SA: rg =0.48, *P* =8.24×10^−32^), but not with OE (OUD: not applicable; SA: rg =0.38, *P* =.092). After conditioning on MDD polygenic risk, the genetic correlation between OUD and SA was still significant (rg=0.40, *P* =3.76×10^−5^).

### Multivariable MR-based association controlling for other psychiatric traits

In the MVMR results involving OUD, SA and those significantly correlated traits (**eTable 8** and **Figure 1F**), genetic liability of SA still retained a significant causal association with OUD (IVW OR= 1.08, 95%CI:1.03-1.13, *P* <0.001), further supported by two additional sensitivity tests (*P* =0.005 and <0.001) and validity checking (*P* >0.05). However, when assessing the genetic liability of OUD with SA, the three MVMR methods did not agree on a consistent causal effect (IVW OR =1.09, 95%CI: 1.00-1.20, *P* =.054). Both patterns were generally stable in repeated MVMR models with reduced number of confounders (**eFigure 5**). A full list of instrumental SNPs involved in MVMR was included in **eTable 9**.

## Discussion

In this study, our triangulated results support more the existence of an MH-independent bidirectional relationship between OUD and SA: 1) the undirected phenotypic and genetic correlations are both significant after adjustment of mental health conditions; 2) from SA to OUD, not only SA polygenicity risk can predict OUD status in opioid-exposed population, but also the genetic liability of SA has a suggestive causal effect on OUD risk; 3) from OUD to SA, though it is underpowered to identify similar causal association, the OUD polygenicity risk can still predict SA status in nonMH population.

To our knowledge, this is the first endeavour using genomic data to explain OUD and SA comorbidity in the context of other mental health conditions. Although observational studies have emphasized the association between substance use disorder (SUD) and suicide idea or behaviours, they were unable to establish causal relationships.^49, 50^ Meanwhile, there are only a few related genetic association studies specifically focusing on correlated relationships between them. Instead, previous studies usually concentrated on SA with psychiatric disorders, especially MDD,^51^ BD,^51, 52^ and SCZ^53^; or they just focused on mental disorders themselves.^32^ Though studies have emphasized that suicidality is genetically independent of major psychiatric diagnoses,^38, 54, 55^ few has unearthed biological mechanism of SA until recent finding from ISGC becoming available.^30^ With similar approaches (e.g., PRS and MR), an earlier report has revealed a potential causal association between self-harm and some complex traits (including mental health disorders and SUD).^17^ Nevertheless, OUD was left behind in their study. Though cross-phenotype genetic correlation and MR were also included in the original MVP OUD GWAS,^31^ it did not consider SA or self-harm in their selected traits. Our findings can fill the knowledge gap remained.

One mechanism could be that impulsiveness that serves in stress-diathesis model might partly account for this special relationship.^56^ Suicide attempters, differ from suicide decedents who are more deliberate, are more linked to impaired probabilistic learning assessed by risk taking tendency^57^ and economic and social reward and punishments,^58, 59^ in another word, are assumed to be of higher trait impulsiveness.^60, 61^ When faced from psychological or material stress, such as stigma, discrimination and violence, they usually lack of social reward, as well as chronically expose to social exclusion. The stress response triggers a psychobiological reaction involving the hypothalamic-pituitary-adrenal axis,^55, 62^ which in turn might potentially exacerbate poor health outcomes and risk behaviours such as opioid abuse.^63^ Another explanation lies on the opioid receptor system. This system may play a role in negative affect associated with suicidality since its abnormality may impact cognition and reward circuitry via altered mu opioid receptor binding,^55, 59^ and buprenorphine, the kappa antagonist to decrease illicit opioid use and reduce the multiple negative consequences of OUD (e.g., fatal and nonfatal substances overdose, infectious complications, and anti-suicidal strengths).^64^ Additional clue comes from our SNP-gene annotation (**eTable S6**) and gene-set enrichment analysis ((**eFigure 4**). Genes mapped by instrumental SNPs included in two-sample MR have not only shown significant effects in impulsiveness trait (e.g., risk-taking tendency and adventurousness) and major psychiatric disorders, but also enriched in metabolic and hematological index abnormalities,^60, 65–68^ indicating their influence in inflammatory and immune dysfunction. This is consistent with pathogenetic mechanism of suicidal behaviours^55^ and suggests a possible risk for OUD and use of anti-inflammatory drugs in both phenotypes.^69^

## Limitations

The main advantage of this study lies on the big genomic data (e.g., UKB, MVP, and ISGC) we can collect for the phenotypes of interest, and the integrated pipeline for cross-phenotype analysis. We also included additional data and sensitivity tests for verifying potential causal associations. Nevertheless, this data-driven study also has some limitations. First, we don’t have access to clinical phenotypes in ISGC SA and MVP OUD studies. Both focused mainly on their primary phenotype and did not especially filter out those participants with the alternative phenotype. Given that OUD is often under diagnosed at population level, there might be hidden OUD patients in ISGC samples that may affect the polygenicity risk of SA. Second, the reason why using opioids, like physical pains of cancer patients, were not taken into account. Hence there might be a horizontal pleiotropy between the pathogenesis of pain and phenotypes (e.g., MDD) we focused on.^32^ Third, this study mainly focused on establishing genetic correlation and causal associations between OUD and SA, but not proceed much to dig up biological mechanisms of their observed comorbidity. Further research should add more biological evidence (e.g., exact genes and pathways) through gene transcription and multi-omics analyses.^70^ Last, this bivariate relationship was based on observational data and statistical approaches. Pragmatic clinical trials and implementation research should be added in the future study before translating it into clinical services.^71^

## Conclusion

In summary, both OUD and SA phenotypes present strong phenotypic and genetic association with each other and major categories of psychiatric diseases. There is also an independent association between OUD and SA after controlling for the influence of psychiatric diseases. Moreover, their relationship is likely to be putatively bidirectional. Comprehensive prevention approaches that address the intersections between suicide, opioid use and shared comorbidity with mental health conditions are needed, so as to provide a promising path forward for addressing these public health challenges among the population.

## Data Availability

All analyses were conducted using publicly available data. Complete genetic datasets for every analysis included in this study are available in the Supplement. The individual-level data publicly available summary-level data is available by application at https://www.ukbiobank.ac.uk/. For summary-level data, OUD and AUD are accessed through NCBI dbGaP for MVP (phs001672.v6.p1)39. MDD, BD, SCZ and OE were accessed at https://www.med.unc.edu/pgc/results-and-downloads/ and through MR Base at https://www.mrbase.org/. Code Availability: the analysis code in R is available on request, and all data displayed in the figures are available in the Supplement.

## Author Contributions

Drs. Qiang Wang Qiang and Hongsheng Gui have full access to all the data in the study and take responsibility for the integrity of the data and the accuracy of the data analyses.

Concept and design: All authors.

Acquisition, analysis, or interpretation of data: Huang, Chen, Wang, Gui.

Drafting of the manuscript: Huang, Wang, Gui.

Critical revision of the manuscript for important intellectual content: All authors.

Statistical analysis: Huang, Chen, Li, Wang, Gui.

Obtained funding: Wang, Gui.

Administrative, technical, or material support: Wang, Gui.

Supervision: Wang, Gui.

## Acknowledgement

This research was made possible by previous studies from PGC, MVP, iPSYCH, the developers of the MRC-IEU UK Biobank, and researchers from ISGC (especially Dr. Niamh Mullins), who provided the latest European-ancestry statistics on SA. We acknowledge their contributing studies and the participants in those studies, without whom this effort would not be possible.

## Software and codes

All analyses were conducted using publicly available data. Complete genetic datasets for every analysis included in this study are available in the Supplement. The individual-level data publicly available summary-level data is available by application at https://www.ukbiobank.ac.uk/. For summary-level data, OUD and AUD are accessed through NCBI dbGaP for MVP (phs001672.v6.p1)^39^. MDD, BD, SCZ and OE were accessed at https://www.med.unc.edu/pgc/results-and-downloads/ and through MR Base at https://www.mrbase.org/. Code Availability: the analysis code in R is available on request, and all data displayed in the figures are available in the Supplement.

