## Supplementary file for "Cross-phenotype relationship between opioid use disorder and suicide attempts: new evidence from polygenic association and Mendelian randomization analyses"

**Huang et al.**

List of material content:

Supplementary methods and results

Supplementary references

Supplementary tables 1-11

Supplementary figures 1-5

**Supplementary methods and results**

Here we provide additional description of data and approaches included in our analytical workflow (**Figure S1**), which was designed for mining of multiple collected datasets in this study (**Table S1**). Part-1 corresponds to data quality control and analyses involving UK Biobank raw phenotypes and genotypes. Part-2 corresponds to technical details for statistical approaches (e.g., Mendelian randomization and genetic correlations) involving only GWAS summary statistics.

**1. UK Biobank data processing and analytical details**

*1.1 Assessment of outcomes and exposures*

Suicide attempts (SA): SA cases were defined by patient-reported answers (data field 20002, 20483, and 40001), ICD 9/10 diagnostic codes (primary and secondary) from in-hospital records and read v2/3 codes in primary care records. Controls were selected from those Mental Health Questionnaire (MHQ) participants with answer “No” to self-harm (data field 20480); in addition, those who overlapped with any SA cases were also excluded.

Opioid use disorder (OUD): OUD cases were defined using either primary and secondary ICD 9/10 diagnoses codes in UKB hospital record, read v2/3 codes in primary care clinical record, and prescription record of three typical OUD treatment medications (methadone, buprenorphine, and naltrexone). To extract matched controls with opioid exposure, the eligible participants need to satisfy three conditions: 1) have prescription records of opioids (Oxycodone, Hydromorphone, Hydrocodone, Fentanyl, Morphine, Tramadol, Codeine, Meperidine, Nalbuphine, Propoxyphene, Butorphanol, Pentazocine, Belladonna, Opium, Levorphanol), 2) not diagnosed with OUD or prescribed with three OUD treatment medications, 3) with >=2 prescription records with time apart for more than 90 days.

For other exposures to be controlled for confounding to either SA or OUD, we included five general categories of psychiatric diseases (depression, anxiety, stress-related disorder, substance misuse [excluding opioid misuse], and psychotic disorders). Psychiatric diseases were defined using corresponding ICD 9/10 codes for each disease according to a previous report on UKB phenotypic data.^1^ Both SA dataset and OUD dataset was then matched against the psychiatric diseases’ datasets. Those SA cases or controls who were diagnosed with any psychiatric disease before were excluded from the overall dataset so as to formulate a stratified dataset, named “SA-noMH”. Similarly, those OUD cases or controls who were diagnosed with any psychiatric disease before were also excluded from overall OUD dataset to formulate another stratified dataset, named “OUD-noMH”.

The detailed diagnostic codes (ICD-9/10, read v2/3), data fields in MHQ, and medication list are shown in **Table S2**.

*1.2 Genotype data and quality control*

After downloading UKB data to our local computing cluster, standard quality assessment and quality control (QA/QC) steps were applied to raw genomic data that covers all participants with SNPs directly genotyped on two Axiom arrays (~50,000 by UK BiLEVE Axiom array and ~450,000 by UK Biobank Axiom array), and SNPs imputed against Haplotype Reference Consortium reference panel.^2,3^ To generate clean dataset for downstream association analysis, we used quality metrics provided by UK Biobank (data field category 100313 that covers genetic ethnic grouping and principal components) and those metrics (e.g., kinship relationship and genotyping quality) generated by KING or PLINK 2.0.^4,5^ In general, we adopted following QA/QC steps from both sample and variant aspects.

- Sample-level: we filtered samples by their self-reported ancestry/ethnicity (data field 21000) to include British White (n=472,152); and then applied category 100313 metrics to include European-ancestry individuals without sex mismatch or low genotyping individuals (n=408,271); lastly, we kept only unrelated samples defined by KING (n=376,906). In the association, only those with non-missing phenotypic information for this study were included, resulting in a total number of 160,418 unique individuals.
- Variant-level: PLINK was used to filter out variants by minor allele frequency (0.01), Hardy-Weinberg equilibrium test (1x10^-6^), genotyping rate (0.95), and imputation info score (0.5). This resulted in ~10M variants. A subset of common SNPs (n= 1028,599) that match with Hapmap3 variants (rsID) were retained for generating polygenic risk scores.

**2. Data harmonization**

For GWAS summary statistics, we extracted all necessary columns (chromosome, position, rsID, alleles, sample size, odds ratio [OR], standard error [SE], and *P*). For those without OR/SE, we computed them by Z-score, sample size, *P*, and minor allele frequency (MAF).^6^ As SA GWAS (ISGC or iPSYCH) provided summary statistics with or without conditioning on psychiatric disorders (mainly MDD), we also created a MDD-conditioned OUD GWAS summary statistics by running mtCOJO^7^ on original MVP OUD and PGC MDD data. Pairwise datasets were harmonized at both sample level and variant level. For PRS, both training and target cohorts were comprised of European-descent, non-overlapping, and unrelated samples. Only common SNPs (MAF >0.05) matched with HapMap3 reference were included. For two-sample MR, only datasets generated from European-ancestry with no or minimum overlapping (<1%) samples were included. Variants were harmonized for their rsID or genomic coordinate, strands, and effect alleles. Variants were also handled to avoid palindrome, with genotypes from 1000 Genome CEU population^8^ as reference.

**3. Technical details for OUD-SA one-sample Mendelian Randomization**

One-sample MR analysis was performed in UK Biobank using individual-level data with participants OUD SNPs, whether diagnosed OUD or self-reported SA without any mental disease diagnosis. One-sample MR was performed by two-stage least square regression among UKB participants, using the polygenic risk score constructed with borderline significant SNPs and 2SMR derived significant SNPs (*P*<5 × 10^-6^) as instrumental variable. The first stage involved regression of OUD (exposure) upon PRS-OUD, with adjustment of age, sex, PC1-4, and array type. In the second stage, the outcome (SA) was then regressed upon the fitted OUD values from the first regression stage. We also investigated the causal effect of the genetic liability of SA upon OUD by reverse-direction one-sample MR analysis. To avoid sample overlapping with UKB, we select iPSYCH SNPs to construct PRS-SA as instrument for SA. The same analyses were repeated using OUD or SA without psychiatric diseases, and instrumental SNPs in OUD or SA GWAS controlling for psychiatric disease status.

**3. Technical details for OUD-SA two-sample Mendelian Randomization**

*3.1 Selection of instrument SNPs*

After variant filtering, the numbers of remained variants were 5,073,203, 4,358,778, 5,904,239 and 8,017,024 for OUD, OUD conditioning on MDD, SA from ISGC (SA and SA conditioning on MDD) and SA from iPSYCH (OUD and OUD conditioning on MDD, BD and SCZ), respectively. Only datasets generated from European-ancestry samples were included, and no overlapping individuals were identified between SA and OUD GWAS. All SNPs were matched to1000 Genome Project CEU population^8^. Instrumental SNPs were selected from those with GWAS p <5×10^−6^ and being uncorrelated (r^2^ < .05). The instrumental SNP numbers were different between IVW and GSMR for difference between harmonization and HEIDI-outlier analysis.

*3.2 Adjustment of confounding factors*

The mtCOJO^7^ was conducted to analyse OUD conditioning on MDD. GWAS of OUD was from MVP, and MDD from PGC. 1000 Genome Project CEU population^8^ was used as reference in the analysis. 4,358,778 SNPs remained to the final statistics.

*3.3 Sensitivity analyses for 2SMR*

Sensitivity analyses for 2SMR involving checking from different aspects:

- On instrumental SNPs: Cochran’s Q statistic was used to quantify the heterogeneity among the selected SNPs. The results of Cochrane Q statistics showed no significant heterogeneity (P = 0.966). The leave-one-out sensitivity analysis was performed to verify the reliability and stability of the causal effect estimates.
- On potential confounders: Bi-directional two-sample MR analysis were performed to investigate causal effects between OUD conditioning on MDD (OUD|MDD) and SA conditioning on MDD (SA|MDD). Instrumental SNPs were selected as having reached selection threshold P-value less than 5 × 10^−6^ and being uncorrelated (r^2^ < 0.05). SA|MDD showed nominal causal relationship to OUD|MDD (OR = 1.135, 95%CI: 1.009, 1.277, *P* = 0.035). MR-Egger (intercept *P* = 0.394) and MR-PRESSO (Global test *P =* 0.605) showed no horizontal pleiotropy.
- On sample overlapping: Bi-directional two-sample MR analysis were performed between OUD (from MVP and OUD|MDD) and SA from iPSYCH Model 1 (contained covariates on gender, years under follow-up where the participant was 15 years of age or older, and the first 10 principal components of genetic ancestry.) and Model 2 (In addition to the covariates used in Model 1, binary covariates for diagnosis of schizophrenia, bipolar disorders, affective disorders, autism spectrum disorders, anorexia, and any other disorder (ICD-10: F chapter) were included.)^9^. Instrumental SNPs were selected by the same threshold as used for MVP OUD and ISGC SA.

*3.4 Annotation for 2SMR instrumental SNPs*

Functional annotation was performed to uncover the likely biological mechanisms linking instrumental SNPs showing causality from both GSMR and twosampleMR. Enrichment for the genes mapped to all (candidate, genes nearest to lead and lead) SNPs in the identified shared loci was evaluated by the Molecular Signatures Database (MsigdB) via a hypergeometric test implemented in FUMA^10^. Genes without unique entrez ID or pathways containing less than 2 genes were removed. The results were adjusted by Benjamini-Hochberg false discovery rate (BH FDR) of 0.05. Results were shown in **Figure S4.**

**4. Technical details for OUD-SA Multivariate Mendelian Randomization**

*4.1 detailed methods of LDSC*

According to the manual of LDSC, the GWAS summary statistics data were first reformatted into the LDSC format using the default parameter by munge_sumstats.py supplied in the LDSC package. Then, the ldsc.py was used to estimate the genetic correlation with the parameter --rg, --ref-ld-chr and --w-ld-chr.

*4.2 detailed methods of MVMR*

For MVMR analyses, we constructed instruments using SNPs in each of the GWASs meeting our single-variable MR selection criteria, described previously. We used the MVMR extension of the inverse-variance weighted MR method and MR-Egger method to correct for both measured and unmeasured pleiotropy. We combined the SNVs from the relevant GWASs: alcohol use disorder, bipolar disorder, major depressive disorder and schizophrenia; and the bidirectional OUD plus SA instruments. In MVMR, we firstly conducted analysis with all the 5 exposures together (Figure 1F, Table S9). Then we performed MVMR using OUD or SA instruments and exposures with significant causality again for validation (Figure S5, Table S10).

**Supplementary Tables**

All 11 tables are included in a separate Excel file.

**Supplementary Figures**

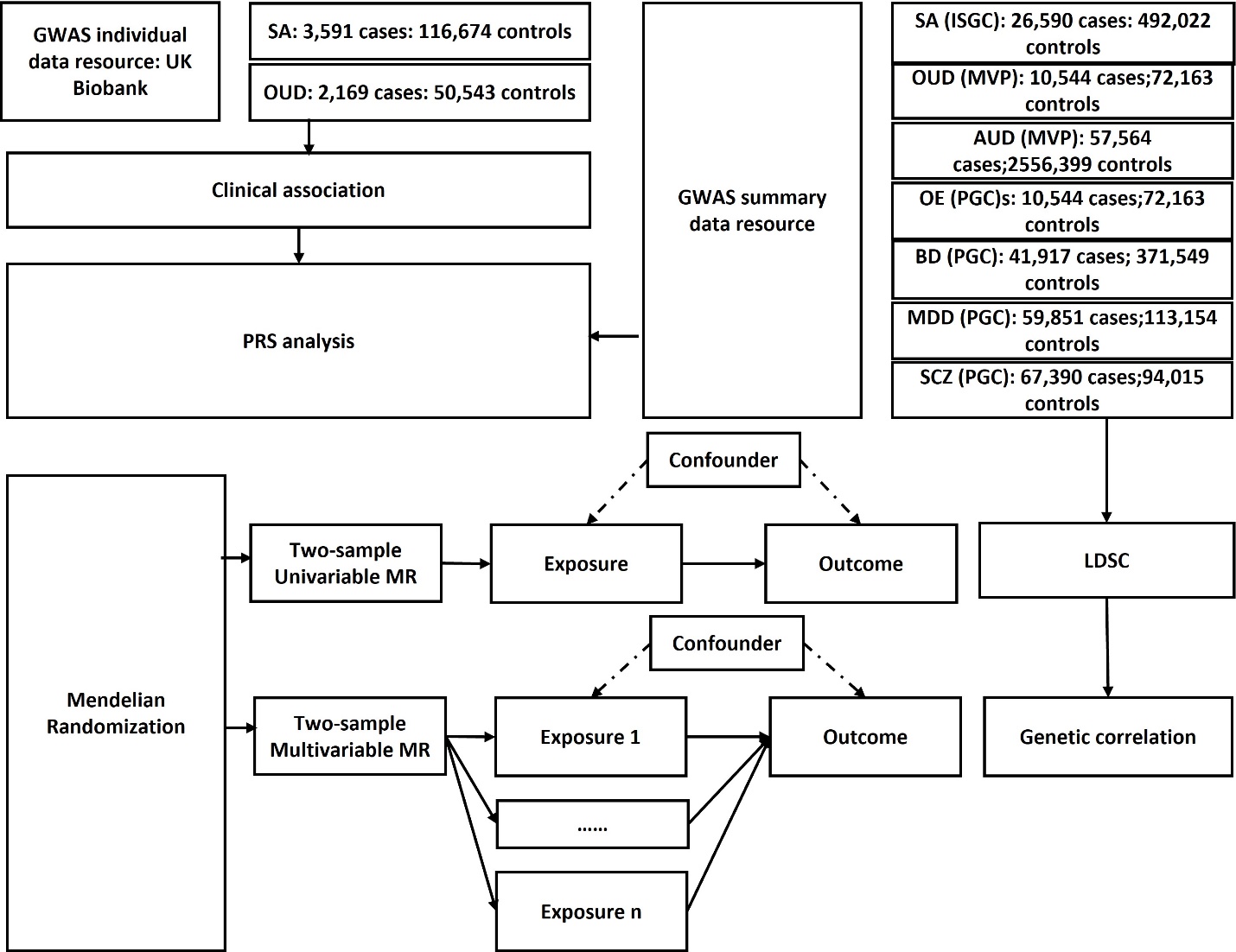

**Figure S1. Workflow of key methodological steps in this study.**

SA: suicide attempt, OUD: opioid use disorder, AUD: alcohol use disorder, OE: opioid exposure; BD: bipolar disorder, SCZ: schizophrenia, MDD: major depression disorder.

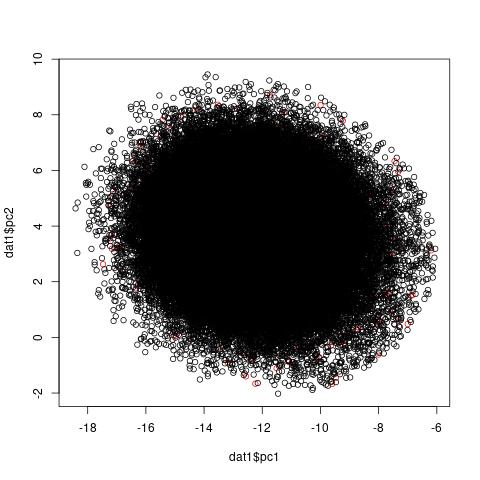

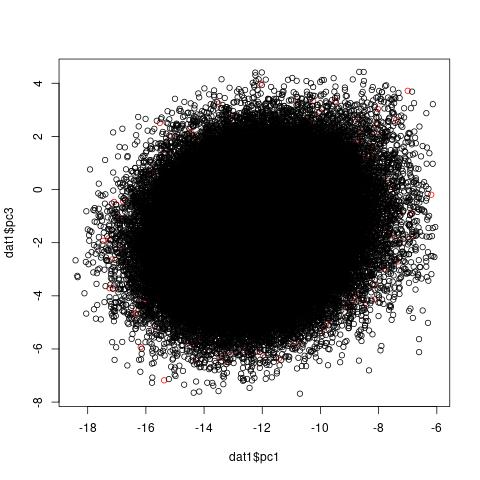

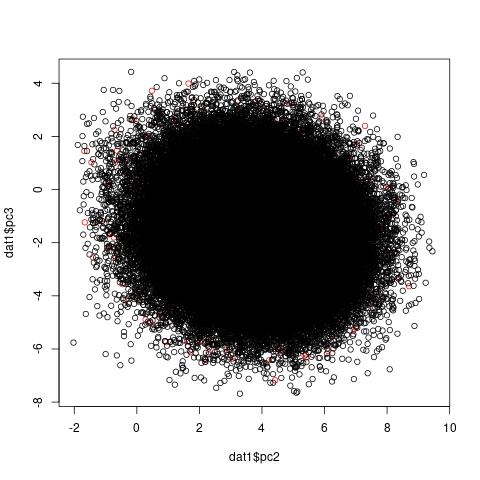
A

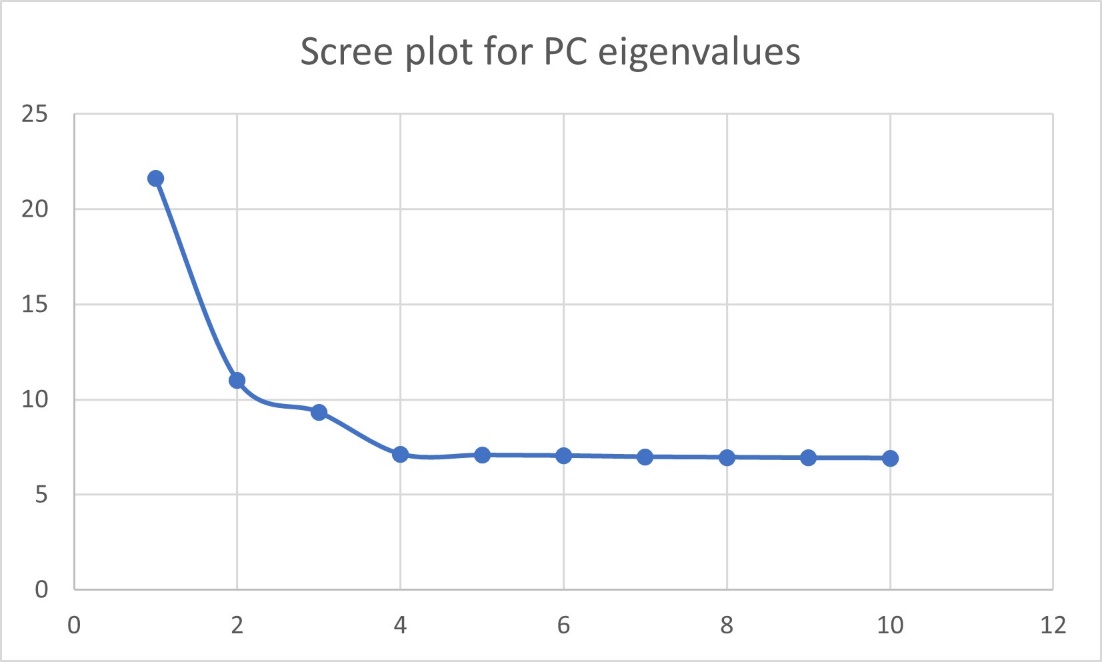
 B

**Figure S2. Principal component analysis in UKB samples.**

A: scatter plot for PC1 vs PC2, PC1 vs PC3, and PC2 vs PC3; B: scree plot for eigenvalues of top 10 PCs.

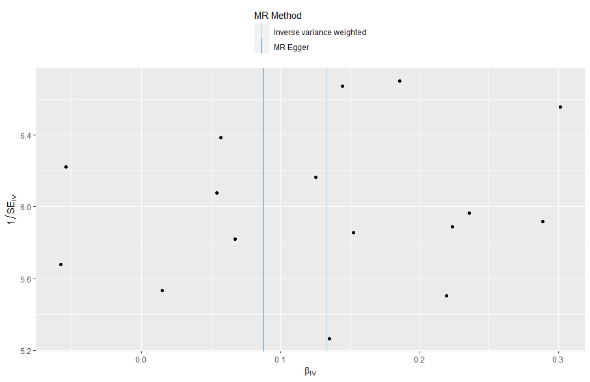

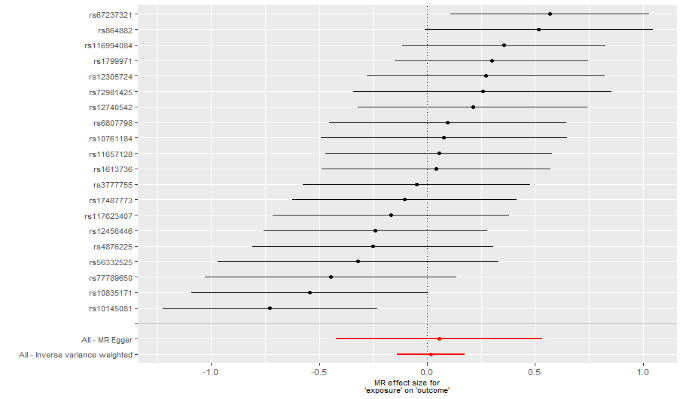

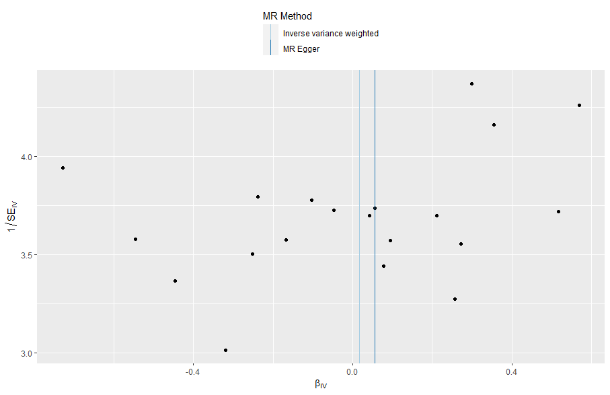

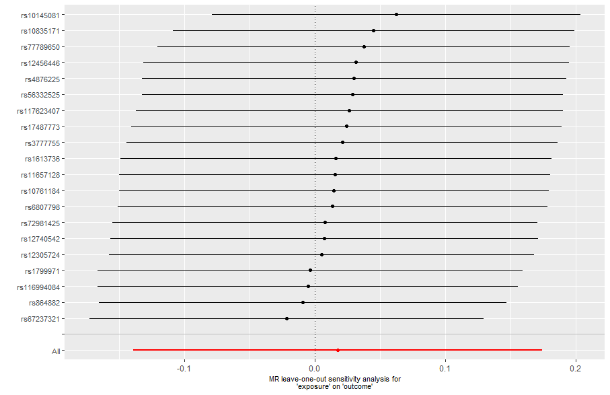

**A)**

**B)**

**D)**

**C)**

**E)**

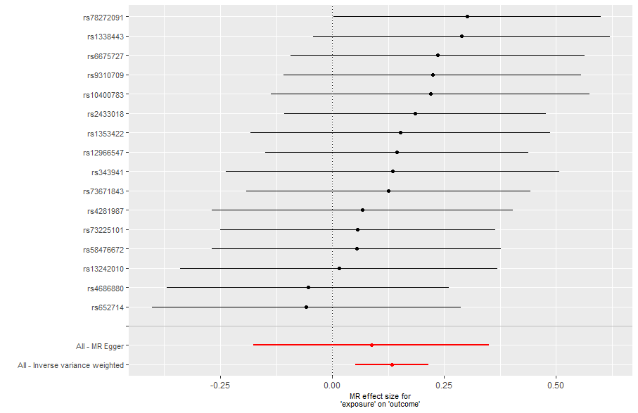

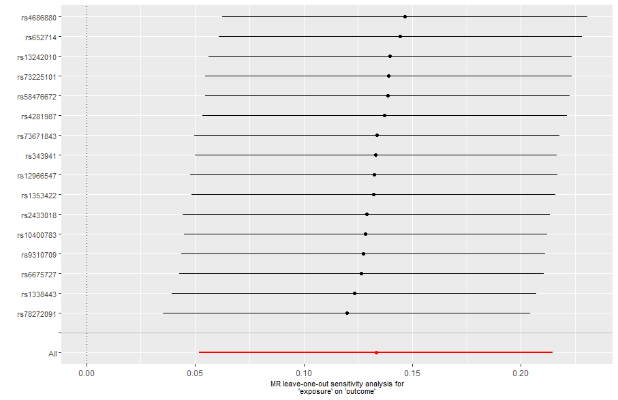

**F)**

**Figure S3. Unsignificant results of Two-sample Mendelian Randomization from OUD to SA and Significant results of Two-sample Mendelian Randomization from SA to OUD..**

A) the forest plot using estimates obtained from individual SNPs. B) the plot showing results from performing leave-one-out analyses. C) The funnel plot for assessing heterogeneity. D) the forest plot using estimates obtained from individual SNPs. D) the plot showing results from performing leave-one-out analyses. F) The funnel plot for assessing heterogeneity.

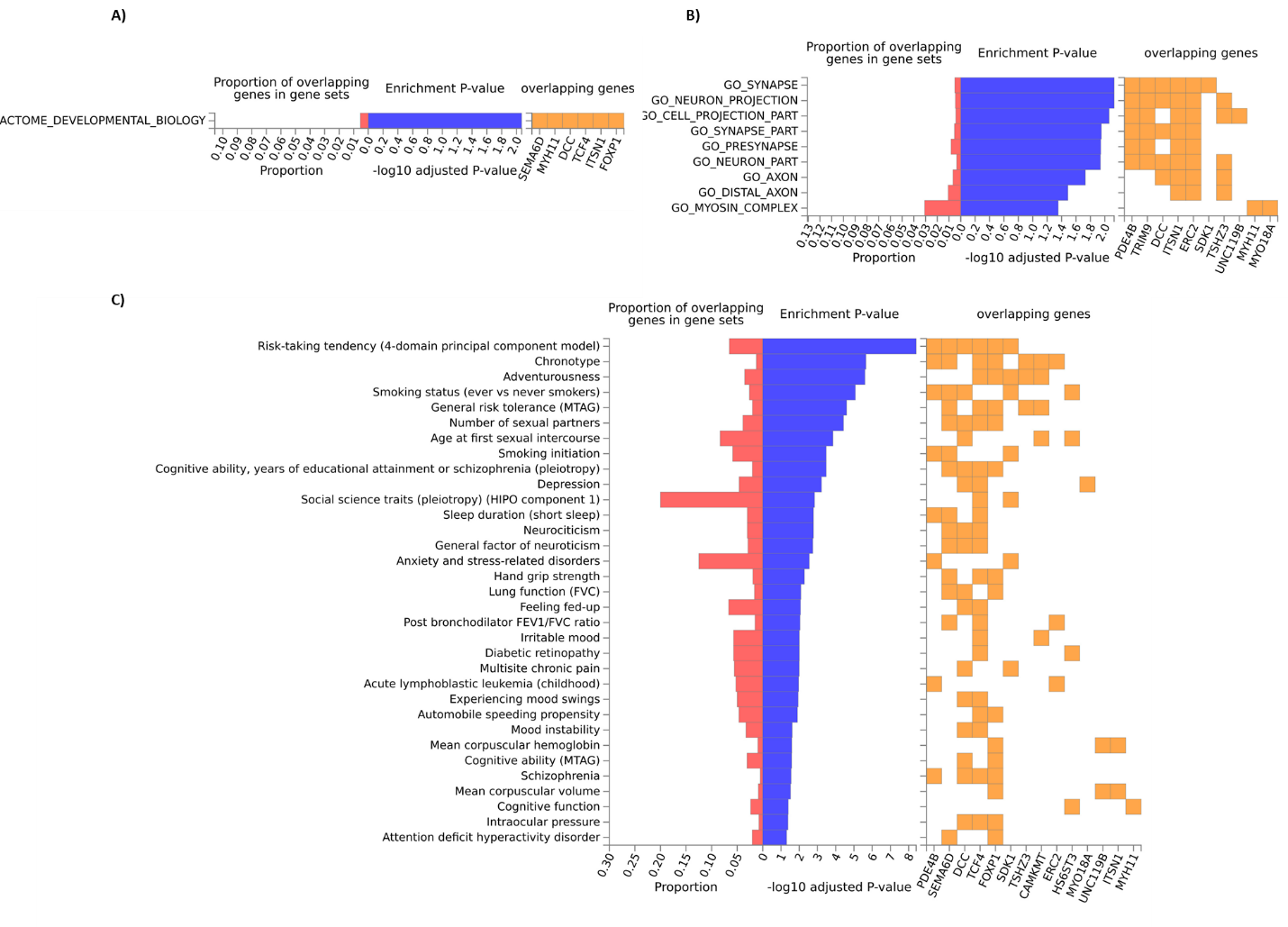

**Figure S4. Significant gene Sets of 2SMR instrumental SNPs.**

A) Gene set of REACTOME. B) Gene sets of GO cellular componets. C) GWAS catalog reported genes.

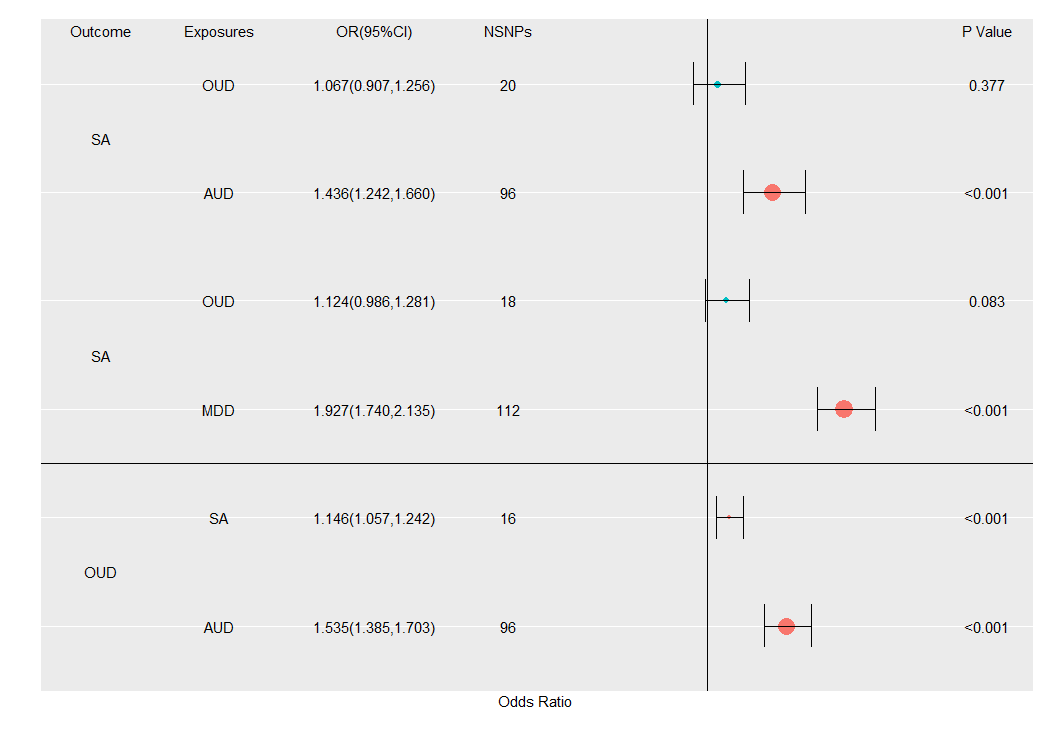

**Figure S5. Odds ratios (ORs) and 95% confidence intervals (CIs) for the effect of SA and OUD and vice versa estimated using the multivariable Mendelian randomization (MR) inverse-variance weighted approach using OUD or SA instruments and exposures with significant causality one by one.**

The vertical line means OR = 1. Red points represent significant causal effects, and green points means unsignificant relationships. The larger point size is, the more significant SNPs of the traits were included in the analyses.
